# Vaccination saves lives: How do patients with chronic diseases and severe COVID-19 fare? Analysis from India’s National Clinical registry for COVID-19

**DOI:** 10.1101/2022.06.22.22276744

**Authors:** Aparna Mukherjee, Gunjan Kumar, Alka Turuk, Ashish Bhalla, Thrilok Chander Bingi, Pankaj Bhardwaj, Tridip Dutta Baruah, Subhasis Mukherjee, Arunansu Talukdar, Yogiraj Ray, Mary John, Janakkumar R Khambholja, Amit H. Patel, Sourin Bhuniya, Rajnish Joshi, Geetha R Menon, Damodar Sahu, Vishnu Vardhan Rao, Balram Bhargava, Samiran Panda

**Affiliations:** Indian Council of Medical Research, New Delhi, India; Postgraduate Institute of Medical Education & Research, Chandigarh, India; Gandhi Medical College, Telangana, India; All Indian Institute of Medical Sciences, Jodhpur, Rajasthan, India; All Indian Institute of Medical Sciences, Raipur Chhattisgarh, India; College of Medicine and Sagore Dutta Hospital, Kolkata, West Bengal, India; Medical College, Kolkata, West Bengal, India; Infectious Disease And Beliaghata Hospital, Kolkata, West Bengal, India; Christian Medical College, Ludhiana, Punjab, India; Smt. NHL, Municipal Medical College, Ahmedabad, Gujarat, India; CIMS Hospital, Ahmedabad, India; All India Institute Of Medical Sciences, Bhubaneswar, India; All India Institute Of Medical Sciences, Bhopal, Madhya Pradesh, India; National Institute of Medical Statistics, Indian Council of Medical Research, Delhi, India; Medstar Health, Baltimore, Maryland, United States of America; National Institute Of Mental Health And Neurosciences, Bangalore, Karnataka, India; Bowring & Lady Curzon Medical College & Research Institute, Bangalore, Karnataka, India; Shaheed Nirmal Mahato Medical College, Dhanbad, Jharkahnd, India; RD Gardi Medical College, Ujjain, Madhya Pradesh, India; Hi Tech Medical College and Hospital, Bhubaneswar, India; Dhiraj Hospital & Sumandeep Vidyapeeth, Vadodara, Ahmedabad, India; Late BRK Memorial Medical College, Jagdalpur, Chhattisgarh, India; Gandhi Medical College, Bhopal, Madhya Pradesh, India; Institute of Medical Sciences & SUM Hospital, Siksha ‘O’ Anusandhan deemed to be University, Bhubaneswar, India; Pandit Bhagwat Dayal Sharma Post Graduate Institute of Medical Sciences, Rohtak, Haryana, India; Government Institute of Medical Sciences, Noida, Uttar Pradesh, India; GMERS Medical College Himmatnagar, Gujarat, India; Gulbarga Institute of Medical Sciences, Kalburagi, Karnataka, India; St. Johns Medical College, Bengaluru, Karnataka, India; S.P.Medical College, Bikaner, Rajasthan, India; SMS Medical College, Jaipur, Rajasthan, India; JN Medical College Aligarh Muslim University, Aligarh, Uttar Pradesh, India; North Eastern Indira Gandhi Regional Institute of Health and Medical Sciences, Shillong, Meghalaya, India; Tata Medical Centre, Kolkata, West Bengal, India; Dr D Y Patil Medical college Hospital and Research centre, Pune, Maharashtra, India; King George Medical University, Lucknow, Uttar Pradesh, India; Mahatma Gandhi Medical College, Jaipur, Rajasthan, India; Kakatiya Medical College, MGM Hospital Warangal, Telangana, India; Department of Health & Family Welfare, Government of Nagaland, Nagaland, India; Community Health Initiative, Nagaland, India; Nizam’s Institute of Medical Sciences, Punjagutta, Hyderabad, India; ESI Hospital and Gayatri Hospital, Raipur, Chhattisgarh, India; Institute of Medical sciences, Banaras Hindu University, Varanasi, Uttar Pradesh, India; ESIC medical College, Sanathnagar, Hyderabad. India; Medanta-The Medicity, Gurugram, Haryana, India; Institute of Postgraduate Medical Education & Research, Kolkata, West Bengal

**Keywords:** SARS-CoV2, mortality, risk factors, outcome, COVID-Vaccine

## Abstract

**Objectives:** This study aims to describe the demographic and clinical profile and ascertain the determinants of outcome among hospitalised COVID-19 adult patients enrolled in the National Clinical Registry for COVID-19 (NCRC).

**Methods:** NCRC is an on-going data collection platform operational in 42 hospitals across India. Data of hospitalized COVID-19 patients enrolled in NCRC between 1^st^ September 2020 to 26^th^ October 2021 were examined.

**Results:** Analysis of 29,509 hospitalised, adult COVID-19 patients [mean (SD) age: 51.1 (16.2) year; male: 18752 (63.6%)] showed that 15678 (53.1%) had at least one comorbidity. Among 25715 (87.1%) symptomatic patients, fever was the commonest symptom (72.3%) followed by shortness of breath (48.9%) and dry cough (45.5%). In-hospital mortality was 14.5% (n=3957). Adjusted odds of dying were significantly higher in age-group ≥60 years, males, with diabetes, chronic kidney diseases, chronic liver disease, malignancy, and tuberculosis, presenting with dyspnea and neurological symptoms. WHO ordinal scale 4 or above at admission carried the highest odds of dying [5.6 (95% CI: 4.6, 7.0)]. Patients receiving one [OR: 0.5 (95% CI: 0.4, 0.7)] or two doses of anti-SARS CoV-2 vaccine [OR: 0.4 (95% CI: 0.3, 0.7)] were protected from in-hospital mortality.

**Conclusions:** WHO ordinal scale at admission is the most important independent predictor for in-hospital death in COVID-19 patients. Anti-SARS-CoV2 vaccination provides significant protection against mortality.

## Introduction

Globally, and in India, the pandemic of SARS-CoV-2 have resulted in unprecedented morbidity and mortality with detrimental effect on healthcare systems and economies. As a response to the pandemic, the ‘National Clinical Registry for COVID-19’ was initiated by the Indian Council of Medical Research (ICMR) in September 2020, with a broad objective to collect good quality, real-time data for evidence-based decision making in clinical practice, public health program and policy.

Hospital mortality among COVID-19 patients has varied from 19-39% across various studies.^1–6^ It has been observed that men, elderly (>60 years of age), and those with comorbidity such as asthma, chronic obstructive pulmonary disease, tuberculosis, pneumonia, diabetes mellitus, hypertension, renal, hepatic, and cardiac diseases and individuals with history of smoking or substance use, history of kidney transplant are at higher risk of developing severe disease or progression to death. The factors associated with such outcomes have been varied; older age being consistent among many populations while others varied amongst studies. ^1–8^

Indian investigations have reported association of old age, presence of diabetes mellitus, presence of severe acute respiratory infection, raised inflammatory markers including interleukin-6, ferritin, lactate dehydrogenase and d-dimer with progression of COVID and/ or related in-hospital mortality^9–12^. Majority of these studies enrolled a small number of participants located at a single centre. Here, we present data from a large cohort of hospitalized COVID-19 patients from 42 hospitals across the country.

The aim of this analysis is to study the demographic profile, clinical characteristics, and outcomes among hospitalised COVID-19 adult patients, enrolled in the National Clinical Registry for COVID-19 (NCRC) and to ascertain the factors associated with predefined outcomes.

## Methods

The National Clinical Registry for COVID-19 (NCRC) is a platform for on-going prospective data collection, developed and maintained by ICMR in collaboration with the Ministry of Health & Family Welfare (MOHFW), Government of India, All India Institute of Medical Sciences, New Delhi (AIIMS) and the ICMR-National Institute of Medical Statistics (NIMS). The structure and protocol of the registry are available in the public domain (https://www.icmr.gov.in/tab1ar1.html). A hub and spoke model has been adopted for this registry. At the beginning, an expression of intent was invited for participation in the registry network; willing hospitals were screened based on a site feasibility matrix. A steering committee with subject experts guides the conduct of the registry and suggests solutions to roadblocks, if any. A monitoring committee consisting of institutional principal investigators oversees the progress of the registry and explores newer ideas and initiatives, to keep the registry dynamic. The central implementation team at ICMR headquarters remains responsible for the overall execution of the project.

Across the network of NCRC, participating hospitals recruited consecutive in-patients, who had COVID-19 infection confirmed by real time-polymerase chain reaction (RT-PCR), Nucleic acid amplification test (NAAT) or Rapid Antigen Test (RAT). Demographic, clinical and outcome data are collected in an on-going manner by the NCRC network. A dedicated team at the respective sites is responsible for data collection and data entry under the supervision of the institutional primary investigator and the central implementation team at ICMR. All the researchers were trained by the central implementation team at ICMR via an online platform. Regular refresher trainings are conducted in order to minimise errors, and to address the gaps created by change of personnel in the teams.

Data is collected using a pre-structured case report form (CRF) and is entered into an electronic portal, which has been developed and is being maintained by the ICMR-NIMS, Delhi. The CRFs include socio-demographic information, symptom and comorbidity profile at the baseline, clinical examination findings at the time of admission and on alternate days during the course of hospital stay, results of laboratory investigations conducted as per treating physician and the outcomes of the hospital stay.

The database platform is hosted on a secure server and is audited by the National Informatics Centre (NIC). Information contained in the database, the configuration of the information within the database, as well as the database itself are fully encrypted. Every client-server data transfer is encrypted through a valid certificate. Data loss is prevented by frequent backup runs.

### Data Analysis

Socio-demographic, clinical, laboratory and hospital outcome data were analysed; categorical data presented as frequency and proportions and continuous data as mean (standard deviation) or median (Inter-quartile range), as appropriate. Logistic regression model was used to determine the factors associated with the outcome of the patients. For the purpose of outcome analysis, death was defined as death due to any cause of a COVID-19 positive patient occurring during hospital stay. Patients who were transferred to another hospital or left against medical advice were excluded from the outcome analyses, though their baseline characteristics were analysed. Age, gender, body mass index, pre-existing comorbidities, lag between symptom onset and admission, laboratory parameters at admission including lactate dehydrogenase (LDH), ferritin, d-dimer, C-reactive protein (CRP) and neutrophil to lymphocyte ratio (NLR), severity assessment by WHO ordinal scale^13^ and status of anti-covid19 vaccination were used as explanatory variables in univariate analysis. Chi Square test, t-test or rank sum test was used to examine the association between explanatory variables and outcome, as appropriate. The variables with significant association and those with known clinical or contextual importance were included in the multivariate logistic regression model. As laboratory values were available for a limited number of participants, separate models were used for each of the biomarkers. Data analysis was carried out using STATA v14 (College Station, TX, US).

### Ethical Aspects

Approval was obtained from the Central Ethics Committee for Human Research at ICMR as well as from the respective Institutional Ethics Committee of each of the participating centres. Considering the observational nature of the registry, and collection of anonymised data being done primarily from the routine case records of the patients, a waiver of consent was granted by the Ethics Committees.

## Results

We present here an analysis of 29,509 hospitalised COVID-19 patients over the age of 18 years who were enrolled in NCRC from 1^st^ September 2020 till 26^th^ October 2021. The mean ± SD age of the study population was 51.1 ± 16.2 years; men (51.1 ± 16 years) being similar to that of women (51 ± 16.5 years). Almost three fourth of the participants were at 40 years of age or above and two-thirds of the study participants in this group were men. The mean ± SD body mass index of the participants was 24.8 ± 4.1 kg/m^2^, with one-third of the participants being within normal range, while over 64% were obese or overweight. (Table 1) Four per cent of the enrolled study participants were health care workers.

**Table 1:**
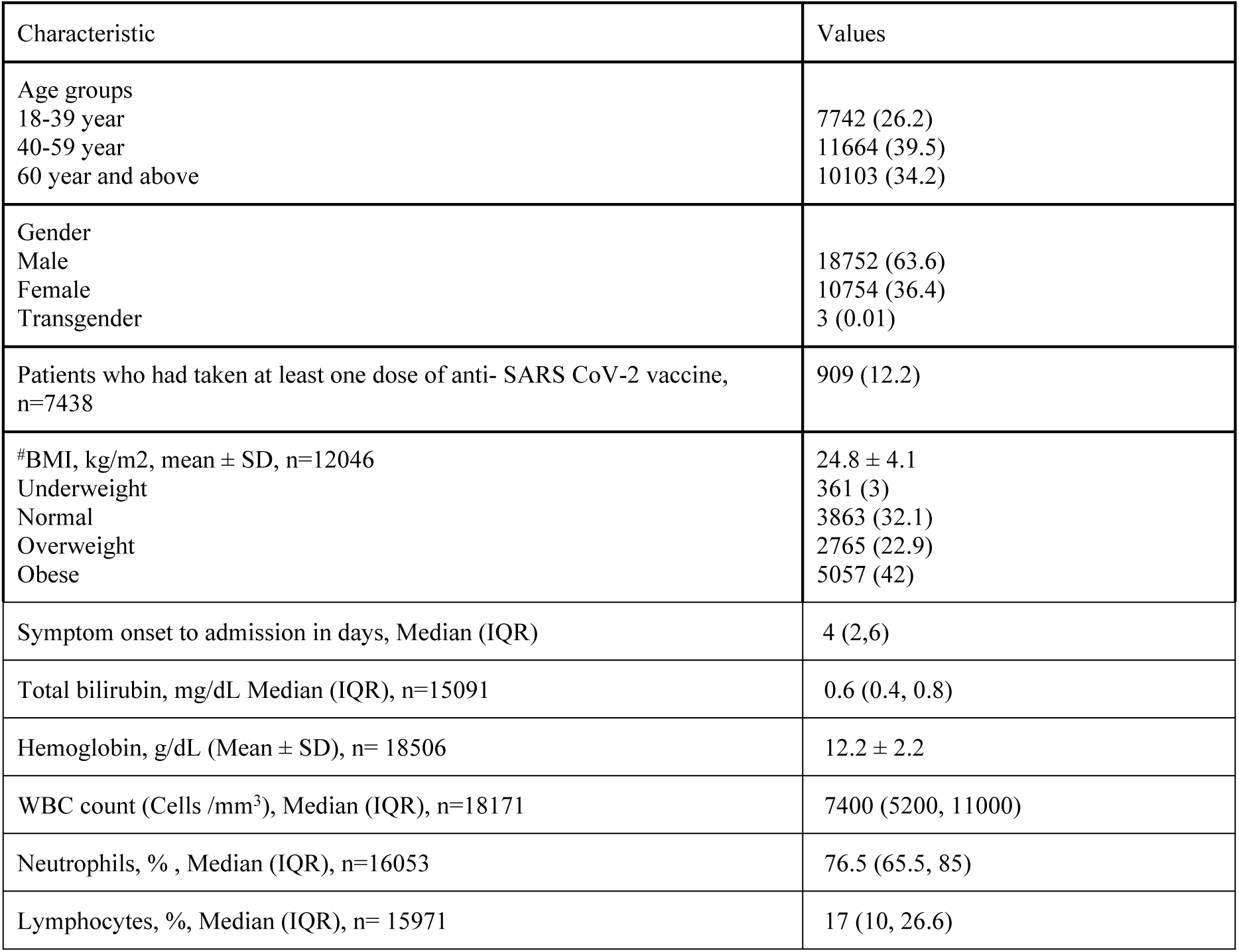
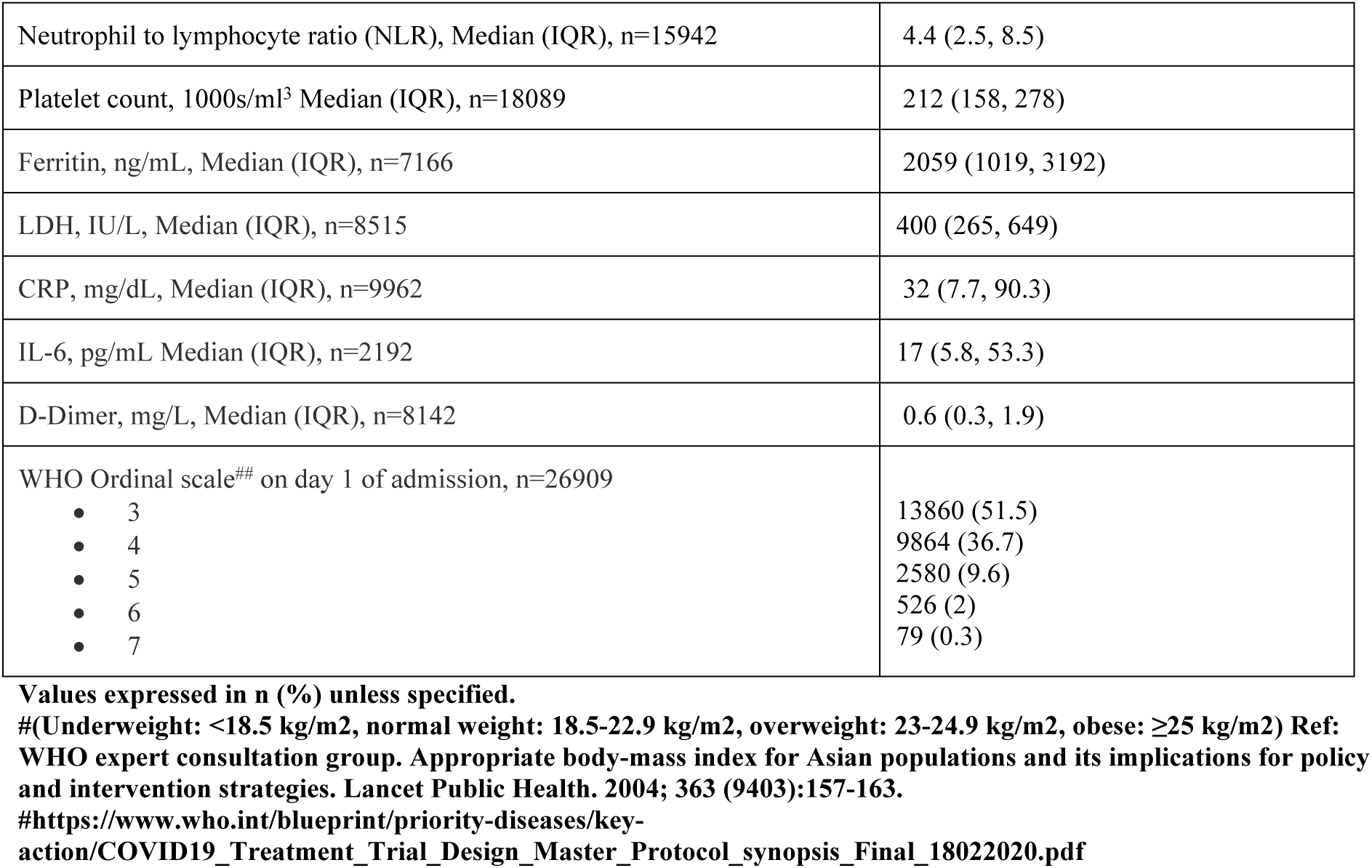
Demographic, symptom and laboratory profile at the time of admission (n=29509)

Of the 29509 patients enrolled, 3794 (12.9%) were asymptomatic at the time of admission and were admitted due to conditions other than COVID-19 and later diagnosed to have COVID-19 or developed COVID-19 during the course of hospitalisation. Among 25715 (87.1%) patients who were admitted with symptoms, fever was the most common symptom (72.3%). Shortness of breath and dry cough was recorded in 48.9% and 45.5% of patients, respectively. Some of the other symptoms were fatigue (20.7%), cough with sputum (14.5%), sore throat (13.5%), muscle ache (12.3%) and headache (11.2%). (S1 Figure)

Median haemoglobin, leucocytes count, neutrophils and lymphocytes were largely within normal limits while the inflammatory markers were raised. (Table 1)

No comorbidities were present in 13831 (46.9%) patients; 15678 (53.1%) participants had at least one comorbidity. Hypertension and diabetes mellitus were the commonest comorbidities reported among 32.4% and 26.2% of patients, respectively. Chronic cardiac disease and chronic kidney disease was present among 5.7% and 3.6% of the study participants, respectively. Other diseases including asthma, malignancy, chronic pulmonary disease, chronic liver disease, stroke, tuberculosis, chronic neurological disease, rheumatologic disease, autoimmune disease, and haematological disorders, HIV infection, Hepatitis B and Hepatitis C infection were each reported in less than 2% of patients. (Figure 1)

**Figure 1:**
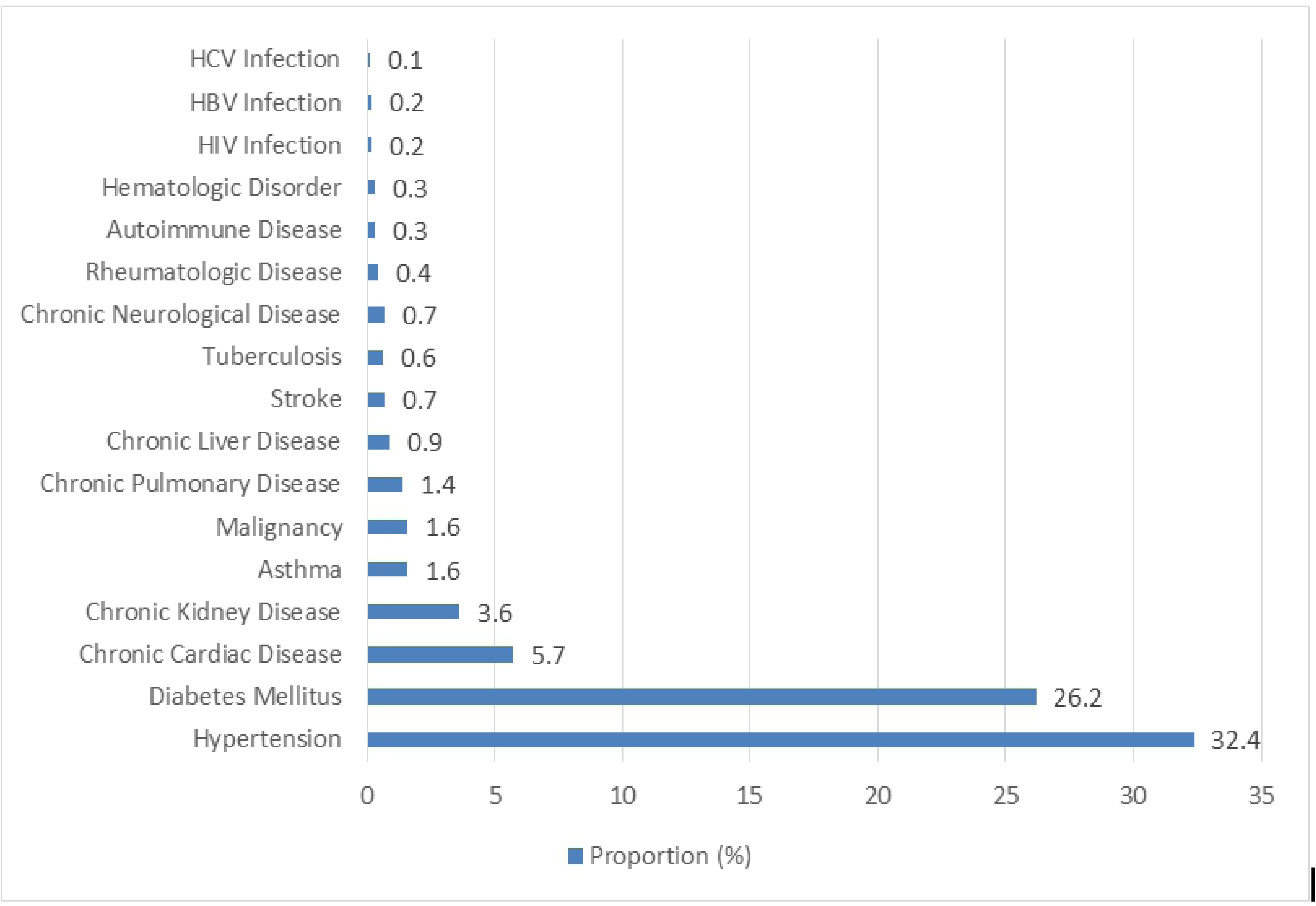
Comorbidity profile of patients, n=29509 HIV: Human Immunodeficiency Virus, HBV: Hepatitis B Virus, HCV: Hepatitis C Virus

The most commonly used drugs were anticoagulants and steroids administered to 60.9% and 60% of patients, respectively. Doxycycline, ivermectin, remdesivir and azithromycin were the other commonly used drugs, while hydroxychloroquine, oseltavimir, faviparavir, IL-6 inhibitors including tocilizumab or itolizumab and convalescent plasma each was administered to less than 5% of patients. More than half of the admitted patients (15922, 54%) required oxygen support during their hospital course, while 2307 (7.8%) required mechanical ventilation. (S1 Table)

Figure 2 shows the monthly trend of selected therapies since the inception of the registry. A marked increase is noticeable in the use of steroid, oxygen supplementation and remdesivir in May 2021, coinciding with the 2nd wave of the pandemic in India dominated by the delta variant of SARS-CoV-2 infection. The use of hydroxychloquine considerably declined after September 2020, while the use of convalescent plasma has been low throughout.

**Figure 2:**
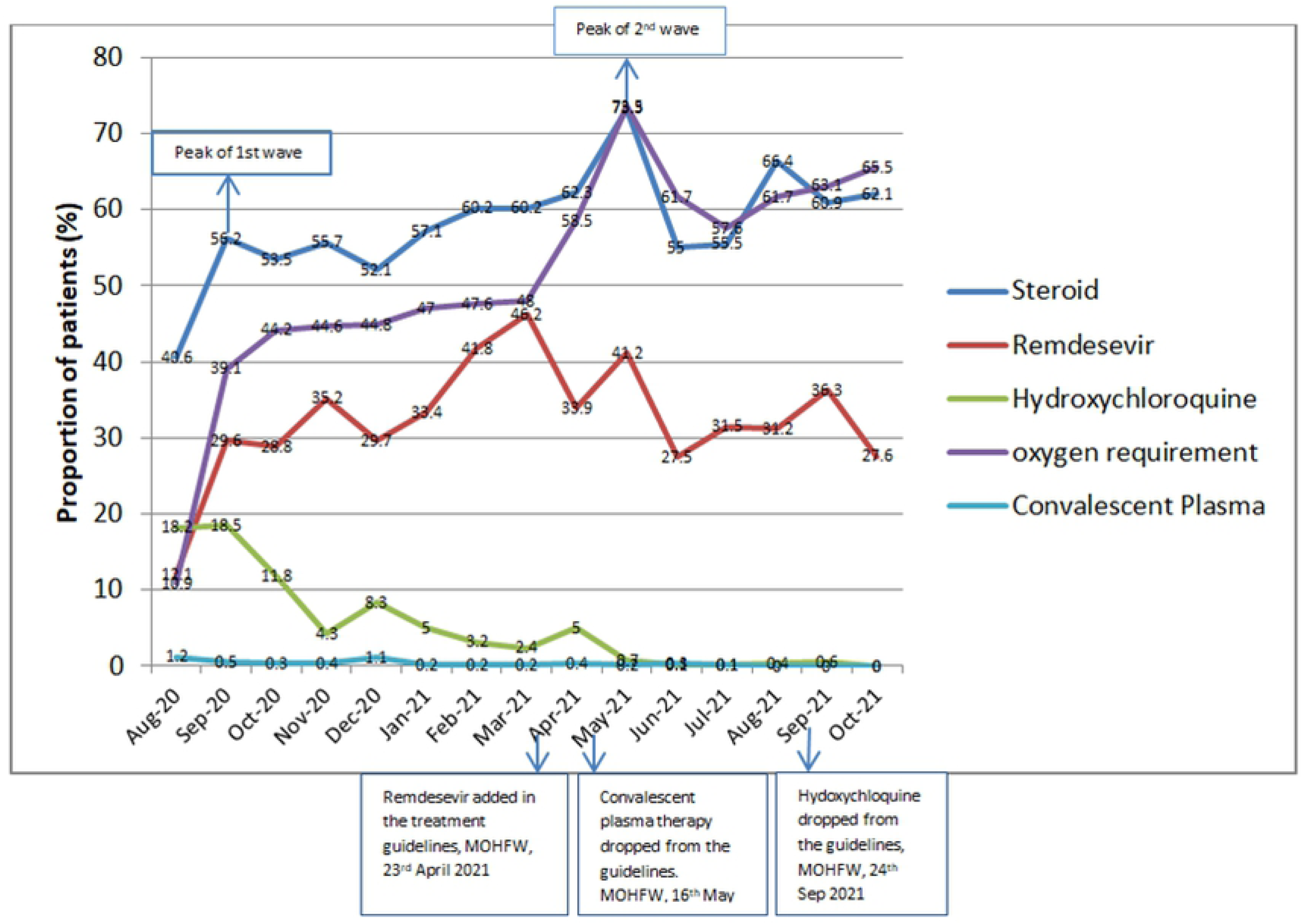
Trends of selected drugs and oxygen requirements

Outcome data on death or discharge were available for 27251 patients; 689 patients who left against medical advice and 1569 patients transferred to other hospitals were excluded from the outcome analysis. In-hospital deaths were reported in 3957 (14.5%) participants and 23294 (85.5%) were discharged. The median duration (IQR) of hospital stay among the study participants was 7 (5, 10) days; 7 (5, 10) days among those discharged and 6 (2, 10) days among those who expired (p= <0.001). Out of the 3957 patients who expired, 3418 (86.4%) died within first 14 days of hospital stay and 539 (13.6%) died after 14 days of hospital admission.

Table 2 shows the association of baseline factors with in-hospital mortality in univariate analysis. Age ≥ 40 year, male gender, comorbidities such as diabetes mellitus, hypertension, chronic cardiac disease, chronic kidney disease, chronic liver disease, malignancy, and stroke, tuberculosis well as respiratory (fast or difficult breathing) or neurological symptoms (altered sensorium or seizures) at presentation and WHO ordinal scale of 4 and above were associated with higher mortality. Receipt of at least one dose of anti-SARS CoV-2 vaccine was associated with lower mortality as compared to the unvaccinated patients [114 (13.3%) vs. 1306 (21.9%), p< 0.001]. Median concentrations of random blood sugar, NLR, LDH, Interleukin-6 (IL-6), CRP, and D-dimer were significantly higher among the patients who died as compared to those who were discharged from hospital. (Table 3)

**Table 2:**
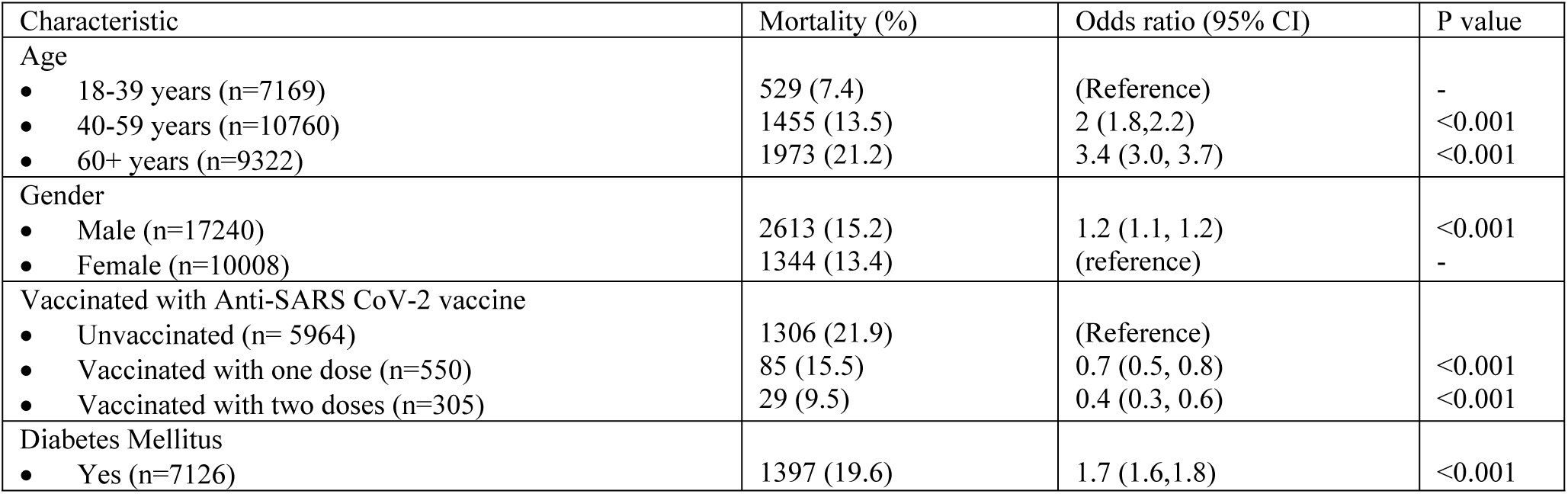
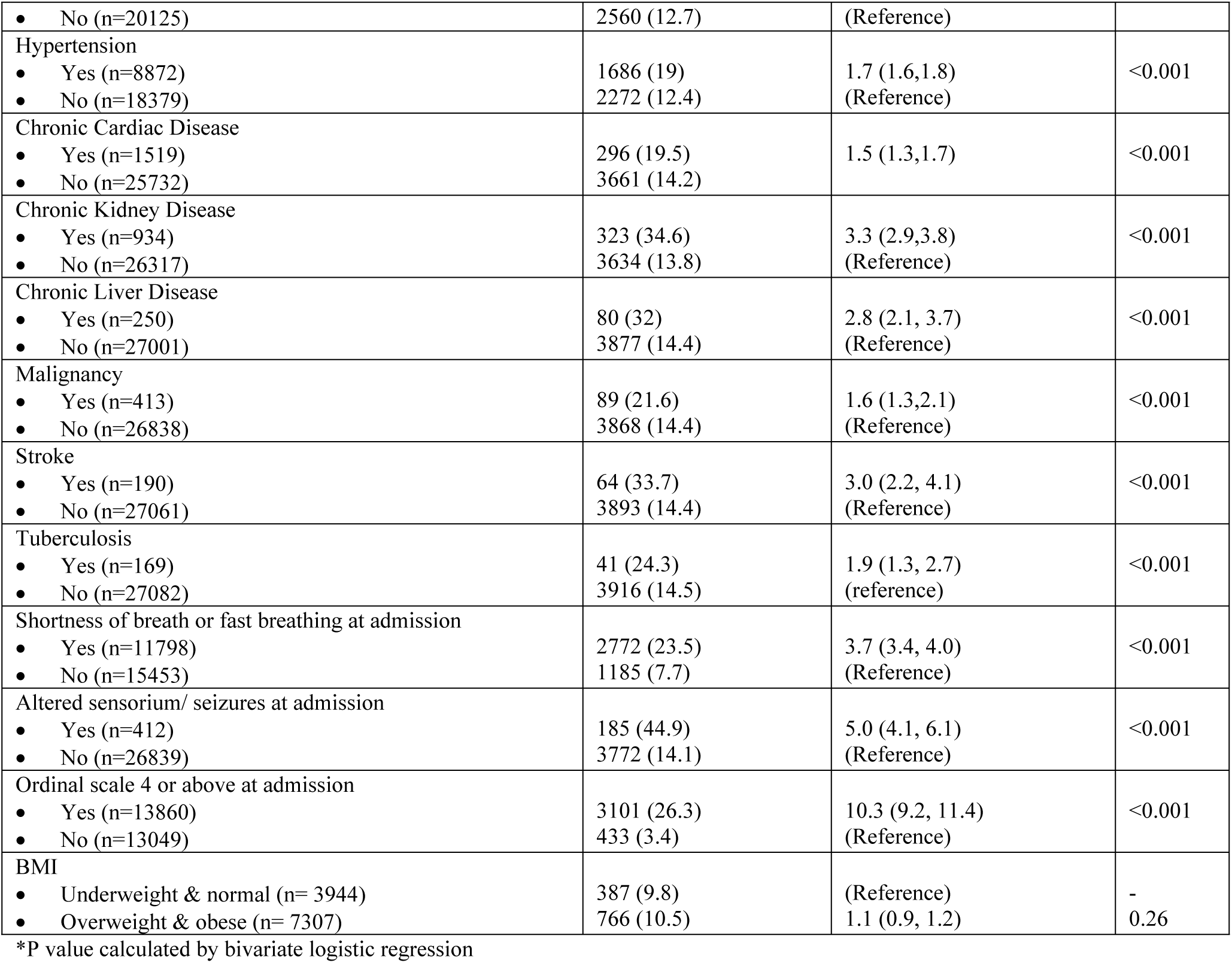
Proportional mortality among hospitalized COVID-19 patients

**Table 3:**
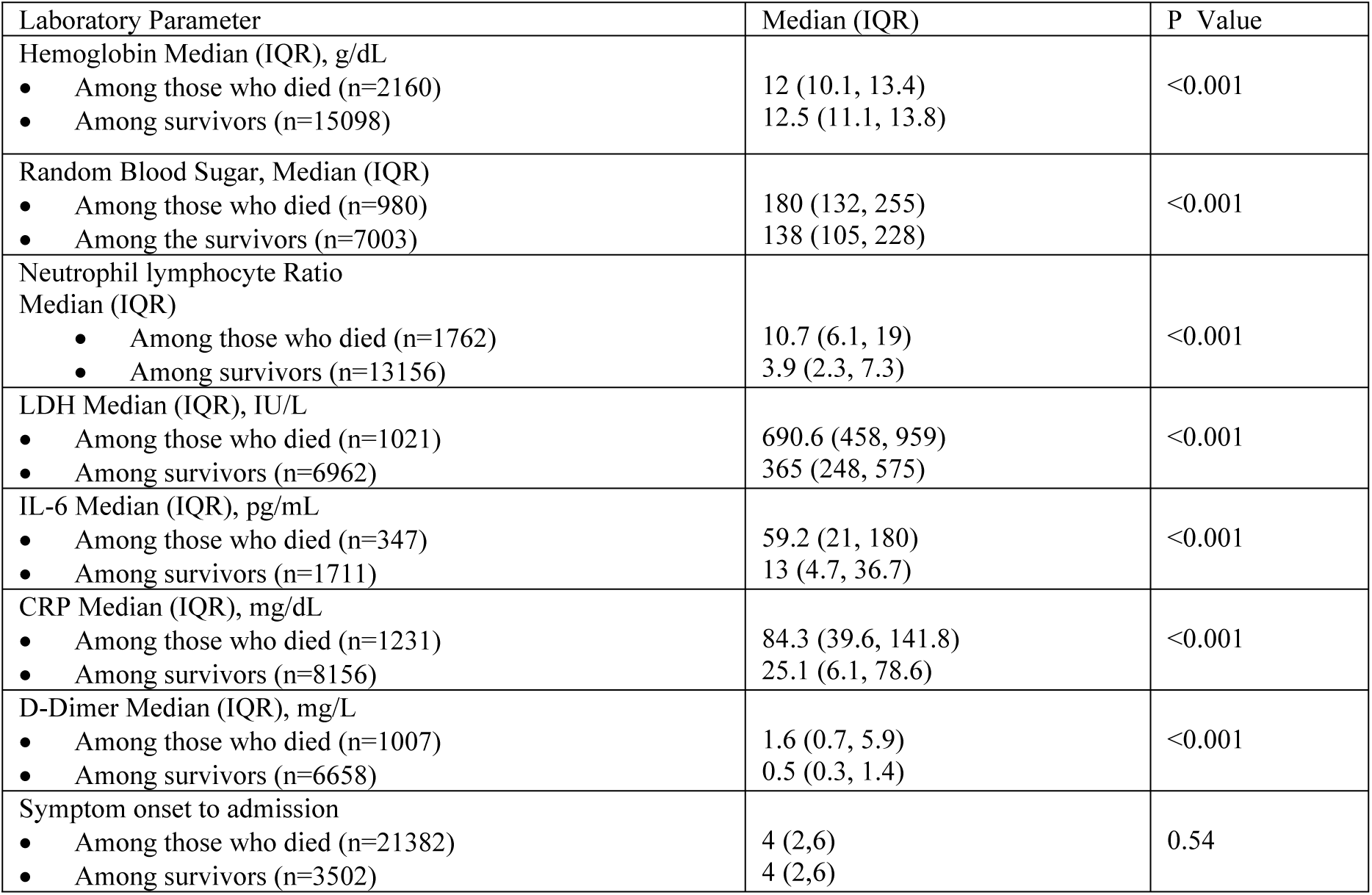
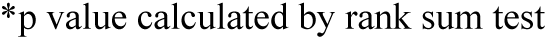
Median laboratory parameters among patients who died and those who survived

| Laboratory Parameter | Median (IQR) | P Value |
| --- | --- | --- |
| Hemoglobin Median (IQR), g/dL |  |  |
| • Among those who died (n=2160) | 12 (10.1, 13.4) | <0.001 |
| • Among survivors (n=15098) | 12.5 (11.1, 13.8) |  |
| Random Blood Sugar, Median (IQR) |  |  |
| • Among those who died (n=980) | 180 (132, 255) | <0.001 |
| • Among the survivors (n=7003) | 138 (105, 228) |  |
| Neutrophil lymphocyte Ratio Median (IQR) |  |  |
| • Among those who died (n=1762) | 10.7 (6.1, 19) | <0.001 |
| • Among survivors (n=13156) | 3.9 (2.3, 7.3) |  |
| LDH Median (IQR), IU/L |  |  |
| • Among those who died (n=1021) | 690.6 (458, 959) | <0.001 |
| • Among survivors (n=6962) | 365 (248, 575) |  |
| IL-6 Median (IQR), pg/mL |  |  |
| • Among those who died (n=347) | 59.2 (21, 180) | <0.001 |
| • Among survivors (n=1711) | 13 (4.7, 36.7) |  |
| CRP Median (IQR), mg/dL |  |  |
| • Among those who died (n=1231) | 84.3 (39.6, 141.8) | <0.001 |
| • Among survivors (n=8156) | 25.1 (6.1, 78.6) |  |
| D-Dimer Median (IQR), mg/L |  |  |
| • Among those who died (n=1007) | 1.6 (0.7, 5.9) | <0.001 |
| • Among survivors (n=6658) | 0.5 (0.3, 1.4) |  |
| Symptom onset to admission |  |  |
| • Among those who died (n=21382) | 4 (2,6) | 0.54 |
| • Among survivors (n=3502) | 4 (2,6) |  |

\*p value calculated by rank sum test

On sub group analysis of patients with diabetes mellitus, malignancy, tuberculosis and those admitted with WHO ordinal scale 4 and above, mortality among vaccinated was significantly lower in each individual subgroup. Among patients with liver disease and kidney disease, though the mortality was lower among the vaccinated but the difference was not statistically significant (Not shown in table).

Factors that were significantly associated with mortality in univariate analysis and those which had clinical relevance were considered in the multivariate logistic models. Odds of in-hospital mortality was significantly and independently higher among patients ≥ 40 year, male gender, with diabetes mellitus, chronic kidney diseases, chronic liver disease, malignancy, and tuberculosis, and those who presented with dyspnoea or neurological symptoms, after being adjusted for other comorbidities such as hypertension, chronic cardiac disease, stroke, severity at admission (WHO ordinal scale), and vaccination status. WHO ordinal scale being 4 and above at admission carried the highest odds of dying [5.6 (95% CI: 4.6, 7.0)]. Patients vaccinated with one and two doses of anti-SARS CoV-2 vaccine had significantly lower odds of dying [OR: 0.5 (95% CI: 0.4, 0.7) for one dose & OR: 0.4 (95% CI: 0.3, 0.7) for two doses]. (Table 4)

**Table 4:**
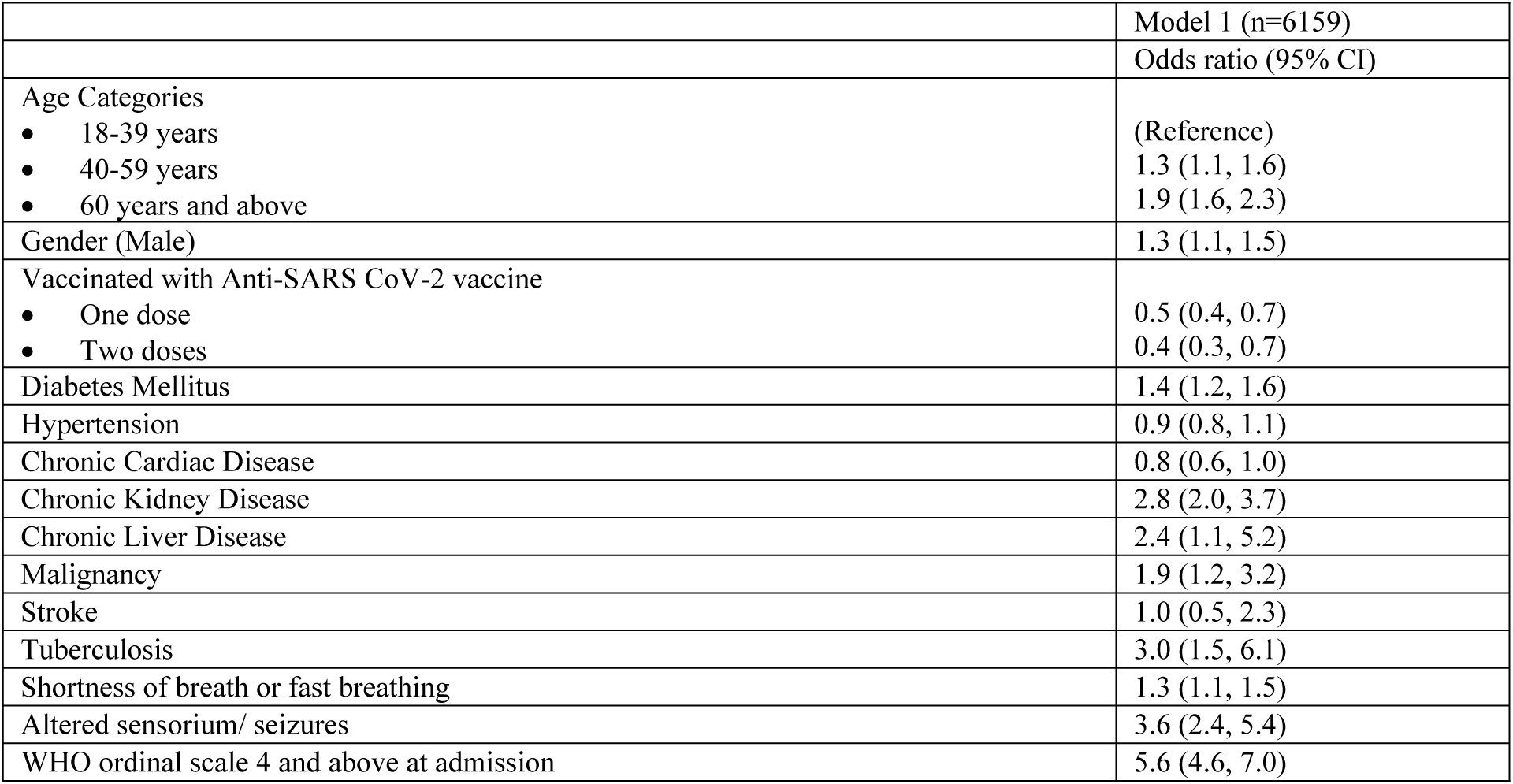
Adjusted Hazards ratio Determinants of hospital deaths using logistic regression

Data available for various baseline laboratory parameters at baseline were limited (as described in table 3). Hence, separate models were tested for each of these parameters. The odds ratio for all biomarkers including haemoglobin, LDH, IL-6, and CRP were statistically significant, but marginally over 1. The odds of death increased by 1.1 for each unit rise in baseline values of NLR and D-dimer separately [OR:1.1 95%CI: 1.1,1.1] after adjusting for age, comorbidities and severity of the illness at admission. The logistic regression models (Model 2 to 8) that included laboratory values are presented as S2 Table. The area under the curve for ROC (AUC ROC) for NLR was 0.79 (95%CI: 0.78, 0.80) and 0.7 (0.68, 0.71). (S2 Figure) Considering the optimal cut-off for NLR as 6.67, both the sensitivity and specificity to classify in-hospital death was 72%; a cut-off of 0.82 mg/L for baseline D-Dimer had a sensitivity and specificity 71% and 65%, respectively.

## Discussion

Our study includes data from 29509 hospitalised COVID-19 patients from 42 hospitals across the country. Apart from the often cited factors such as diabetes mellitus, male gender and advanced age, our study highlighted the association of other comorbidities such as chronic kidney disease, chronic liver disease, malignancy and tuberculosis with increased in-hospital mortality of COVID-19 patients in Indian settings. The importance of anti-SARS-CoV-2 vaccination in protecting against mortality was also evident from our analysis.

More than half of our study participants had at least one co-morbidity, most common being hypertension & diabetes mellitus. The proportion of COVID-19 in-patients having hypertension is similar to the overall population level frequency of hypertension recorded among adult Indians. On the contrary proportion diabetics seem to be much higher in this cohort than the national average.^13,14,15^ Multiple studies have confirmed that following SARS-CoV-2 infection, diabetics are more likely to be hospitalised as compared to non-diabetics, especially if there is poor glycaemic control. ^16^ Diabetes causes an inhibition in neutrophil chemotaxis, phagocytosis, and intracellular destruction of microbes, thus offering efficient virus entry and decreased viral clearance.^17^

In our study, patients above 40 years of age had 1.3 times higher adjusted odds of dying than the younger patients, which increased to 2.1 time with advanced age ≥ 60 year. Advanced age, especially ≥ 60 year, is an established independent risk factor for dying in COVID-19 patients, as shown in various studies across multiple countries since the onset of the pandemic.^18,15^ However, the working age population above 40 years of age also have been deeply affected as shown in our investigation. It would be prudent to include them in all preventive measures and triaging strategies for severity. Age-standardized mortalities for COVID-19 in India, analysed from the Integrated Disease Surveillance Programme special surveillance data, showed that along with the elderly above 60 years of age, the age group of 45-59 years were also affected. ^20^ This could be partly explained by the fact that forced expiratory volume is generally seen to decline after the age of 30-40 years.^21^ Additionally, older age group is known to have a higher prevalence of chronic diseases, which could further attenuate the already dysregulated immune response.

Studies from various parts of India and the world have reported co-morbidities in various combinations to be associated with in-patient mortality in COVID-19 patients.^16,173,24,25,26^. Along with the more recognized risk factors such as diabetes mellitus, chronic kidney disease, and malignancy, our investigation unearthed the independent association of chronic liver disease with higher odds of dying among COVID-19 in-patients. Another retrospective analysis from a single centre in South India linked chronic liver disease with in-hospital mortality of Covid-19 patients.^27^ Increased systemic inflammation, immune dysfunction, coagulopathy and intestine dysbiosis are the hypothesised mechanisms. Important to note in this context is that, the pandemic has been associated with poor eating habits and increased alcohol intake, which might lead to an increase in severity of liver diseases.^8,18,19,29^

Presence of tuberculosis (on-treatment TB) was an important factor associated with higher in-hospital mortality in our cohort. This finding carries a significant relevance in a high TB burden country like India. Increased severity and mortality have been reported in COVID-19 patients with tuberculosis in two metanalyses which included only 26 and 34 Indian patients, respectively ^30,20^. Our study provides more robust supportive evidence for association of present tuberculosis status withCOVID-19 mortality.

Severity of illness at admission as evident by presenting complaints of respiratory or neurological symptoms and WHO ordinal scale 4 or above had higher odds of dying in our registry participants. Similar observations have been reported from a few single centre studies from India, where majority of non-survivors required early oxygen supplementation (oxygen requirement is at WHO ordinal scale 4 and above) ^22,23^.

The baseline laboratory markers including neutrophil-lymphocyte ratio (NLR), LDH, D-Dimer, IL-6 and CRP were higher among the non-survivors, though on multivariate analysis, the odds ratio was marginally above one with minimal clinical relevance, except for NLR and D-dimer. Previous studies have shown that raised biomarkers such as IL-6, CRP, LDH, and NLR are associated with higher mortality.^32-35^ Considering these markers are non-specific indicators of inflammation, the baseline values of Il-6, CRP or LDH seem to offer limited benefit in meaningful prediction of mortality. NLR being a readily available marker can be used to prognosticate outcomes at admission as can D-Dimer. However, guidelines for clinical management from other countries have also stated that there is no consensus in the evidence supporting use of any of the inflammatory markers or D-dimer at baseline to stratify the risk and decide therapeutics. ^36^

Importantly, the current study underlined the protection provided by COVID-19 vaccination against in-hospital mortality. COVID-19 vaccine, irrespective its type, reduced the odds of dying by 50% with one dose and by 60% with two doses. Other smaller, single centre studies from both South and North India have demonstrated the effectiveness of COVID-19 vaccination in reducing mortality.^37,38^ These findings along with mathematical modelling based projections^39^ underscore the key role of vaccine in mitigating the impact of COVID pandemic and managing the burden it poses on the healthcare system.

### Limitations

As this was a record-based study in hospitals maintaining paper-based records, the identification of symptoms, comorbidities, complications and laboratory parameters relied on the accuracy of the records maintained. Secondly, the patients who were transferred to other institutes or who had left against medical advice were not followed up and could not be included in mortality analysis as their outcomes were unknown.

### Strengths

The current analysis from the National Clinical Registry for COVID-19, to the best of our knowledge, is the largest widely representative to examine the association of demographic, clinical characteristics, and laboratory parameters with mortality among hospitalised COVID-19 patients in India. This data captures information from various geographical zones of India involving multiple centres.

## Conclusion

The current investigation highlights the importance of age ≥ 40 years and comorbidities like chronic liver disease, and tuberculosis as predictors of in-patient mortality along with the oft-reported risk factors such as male gender, diabetes mellitus, chronic kidney disease, and baseline severity of illness. WHO ordinal scale 4 and above was an important independent factor associated with in-patient mortality. Interestingly, the baseline values of CRP, IL-6 and LDH offered little help in predicting the outcome, though NLR and D-dimer can be used to classify in-hospital outcomes with a sensitivity and specificity ranging from 65% to 72%. On an encouraging note, vaccination against COVID-19 clearly lowered the risk of dying from the disease and featured as an important armamentarium in our fight against COVID-19 pandemic.

## Data Availability

As the project is currently ongoing, the data is not being shared. Though data can be shared, with a formal proposal sent to the corresponding author.

## Author statements

### Author Contributions

SP is the guarantor. Study design, data analysis, data interpretation and manuscript writing team: AM, GK, AT, AB, TCB, PB, TDB, SM, AT, YR, MJ, JRK, AHP, SB, RJ, GRM, DS, VVR, BB, SP, AA. Monitoring and Conduct of the study: AM, GK, AT, LKS, PM, YP. Patient enrolment, conduct of study, clinical care and data collection: AB, TCB, PB, TDB, SM, AT, YR, MJ, JRK, AHP, SB, RJ, GDP, VS, KS, RM, VSA, MAM, DK, SS, SM, PKK, AK, AS, AP, SC, MD, TM, SC, BB, SRP, DM, SC, AA, DV, MT, NS, MP, SM, AD, KYL, MR, CGS, UKO, RRJ, AK, AP, AS, MP, LS, MR, ADS, LK, PP, ND, SD, JS, AM, LP, JPS, SS, VKK, AK, NY, RU, SS, AS, NNS, NMS, KR, HP, PRM, MKP, SS, AK, MP, MA, DP, VS, SA, RC, MR, ND, BKG, BK, JG, SB, AA, MS, NF, SP, VN, SC, SM, SKS, ST, PL, HD, AG, VK, NS, RV, AP, MPK, ABR, NK, RK, KM, YSR, AM, JC, MC, RKB, MAM, SK, PS, SG, AH.

---

S1 Figure: Clinical characteristics of the study subjects at the baseline among the symptomatic patients, n=25715

S2aFigure: ROC for baseline neutrophil-lymphocyte ratio classifying in-hospital outcomes.

S2bFigure: ROC for baseline D-Dimer levels classifying in-hospital outcomes.

S1 Table: Treatment profile during hospital stay, n=29509

S2 Table: Adjusted odds ratio for determinants of hospital deaths among hospitalised COVID-19 patients using logistic regression model

S1 File: supplement

## Bibliography

1. Becerra-Muñoz VM, Núñez-Gil IJ, Eid CM, García Aguado M, Romero R, Huang J, et al. Clinical profile and predictors of in-hospital mortality among older patients hospitalised for COVID-19. Age Ageing. 2021 Feb 26;50(2):326–34.

2. Rosenthal N, Cao Z, Gundrum J, Sianis J, Safo S. Risk Factors Associated With In-Hospital Mortality in a US National Sample of Patients With COVID-19. JAMA Netw Open. 2020 Dec 1;3(12):e2029058.

3. Gray WK, Navaratnam AV, Day J, Babu P, Mackinnon S, Adelaja I, et al. Variability in COVID-19 in-hospital mortality rates between national health service trusts and regions in England: A national observational study for the Getting It Right First Time Programme. EClinicalMedicine. 2021 May;35:100859.

4. Khamis F, Memish Z, Bahrani MA, Dowaiki SA, Pandak N, Bolushi ZA, et al. Prevalence and predictors of in-hospital mortality of patients hospitalized with COVID-19 infection. J Infect Public Health. 2021 Jun;14(6):759–65.

5. Ruscica M, Macchi C, Iodice S, Tersalvi G, Rota I, Ghidini S, et al. Prognostic parameters of in-hospital mortality in COVID-19 patients—An Italian experience. Eur J Clin Invest. 2021 Sep;51(9).

6. Sarfaraz S, Shaikh Q, Saleem SG, Rahim A, Herekar FF, Junejo S, et al. (2021) Determinants of in-hospital mortality in COVID-19; a prospective cohort study from Pakistan. PLoS ONE 16(5): e0251754. https://doi.org/10.1371/journal.pone.0251754.

7. Galiero R, Pafundi PC, Simeon V, Rinaldi L, Perrella A, Vetrano E, et al. Impact of chronic liver disease upon admission on COVID-19 in-hospital mortality: Findings from COVOCA study. PloS One. 2020;15(12):e0243700.

8. Iavarone M, D’Ambrosio R, Soria A, Triolo M, Pugliese N, Del Poggio P, et al. High rates of 30-day mortality in patients with cirrhosis and COVID-19. J Hepatol. 2020 Nov;73(5):1063–71.

9. Mammen JJ, Kumar S, Thomas L, Kumar G, Zachariah A, Jeyaseelan L, et al. Factors associated with mortality among moderate and severe patients with COVID-19 in India: a secondary analysis of a randomised controlled trial. BMJ Open. 2021 Oct 4;11(10):e050571.

10. Zodpey SP, Negandhi H, Kamal VK, Bhatnagar T, Ganeshkumar P, Athavale A, et al. (2021) Determinants of severity among hospitalised COVID-19 patients: Hospital-based case-control study, India, 2020. PLoS ONE 16(12): e0261529. https://doi.org/10.1371/journal.pone.0261529.

11. Agarwal. Epidemiological determinants of COVID-19 infection and mortality: A study among patients presenting with severe acute respiratory illness during the pandemic in Bihar, India [Internet]. [cited 2022 Jan 17]. Available from: https://www.npmj.org/article.asp?issn=1117-1936;year=2020;volume=27;issue=4;spage=293;epage=301;aulast=Agarwal

12. Mishra V, Burma AD, Das SK, Parivallal MB, Amudhan S, Rao GN. COVID-19-Hospitalized Patients in Karnataka: Survival and Stay Characteristics. Indian J Public Health. 2020 Jun;64(Supplement):S221–4.

13. Ramakrishnan S, Zachariah G, Gupta K, Shivkumar Rao J, Mohanan PP, Venugopal K, et al. Prevalence of hypertension among Indian adults: Results from the great India blood pressure survey. Indian Heart J. 2019 Jul 1;71(4):309–13.

14. Anjana RM, Deepa M, Pradeepa R, Mahanta J, Narain K, Das HK, et al. Prevalence of diabetes and prediabetes in 15 states of India: results from the ICMR-INDIAB population-based cross-sectional study. Lancet Diabetes Endocrinol. 2017 Aug;5(8):585–96.

15. O’Driscoll M, Ribeiro Dos Santos G, Wang L, Cummings DAT, Azman AS, Paireau J, et al. Age-specific mortality and immunity patterns of SARS-CoV-2. Nature. 2021 Feb;590(7844):140–5.

16. Barman Roy D, Gupta V, Tomar S, Gupta G, Biswas A, Ranjan P, et al. Epidemiology and Risk Factors of COVID-19-Related Mortality. Cureus. 13(12):e20072.

17. Padmaprakash KV, Vardhan V, Thareja S, Muthukrishnan J, Raman N, Ashta KK, et al. Clinical characteristics and clinical predictors of mortality in hospitalised patients of COVID 19 : An Indian study. Med J Armed Forces India. 2021 Jul;77(Suppl 2):S319–32.

18. Wang Q, Davis PB, Xu R. COVID-19 risk, disparities and outcomes in patients with chronic liver disease in the United States. EClinicalMedicine. 2020 Dec 22;31:100688.

19. Marjot T, Webb GJ, Barritt AS, Moon AM, Stamataki Z, Wong VW, et al. COVID-19 and liver disease: mechanistic and clinical perspectives. Nat Rev Gastroenterol Hepatol. 2021 Mar 10;1–17.

20. Gao Y, Liu M, Chen Y, Shi S, Geng J, Tian J. Association between tuberculosis and COVID-19 severity and mortality: A rapid systematic review and meta-analysis. J Med Virol. 2020 Jul 28;10.1002/jmv.26311.

